# Rapid displacement of SARS-CoV-2 variants within Japan correlates with cycle threshold values on routine RT-PCR testing

**DOI:** 10.1101/2022.04.13.22273855

**Authors:** Danelle Wright, Carmen Chan, Wirawit Chaochaisit, Mio Ogawa, Junko Tanaka, Satoshi Nozaki, Shinji Narita, Eisuke Shimizu, Hideyuki Aoshima, Iri Sato Baran

**Author notes:** **Corresponding Author** Danelle Wright, Genesis Institute of Genetic Research, Genesis Healthcare Corporation, Yebisu Garden Place Tower 26F 4-20-3 Ebisu, Shibuya-ku, Tokyo, Japan.

## Abstract

**Background:** The rapid spread of SARS-CoV-2 worldwide has led to the emergence of new variants due to the presence of mutations that alter viral characteristics, but there have been few studies on trends in viral lineages in Japan, an island country. We hypothesized that changes in cycle threshold (Ct) values on reverse transcription polymerase chain reaction (RT-PCR) reflect the prevalent variants during a given period.

**Methods:** We performed next-generation sequencing of positive samples to identify the viral lineages in Japan in 2021 and compared variant prevalence with the average Ct values on routine RT-PCR using 4 primer sets.

**Results:** Based on 3 sequencing runs, the highly transmissible Alpha variant, which prevailed over other lineages, such as R.1, from April 2021, was dominated by the even stronger Delta variant between July and August 2021 in Japan. The decrease in our routine RT-PCR Ct values with 4 primer sets correlated with these fluctuations in lineage prevalence over time.

**Conclusions:** We confirmed that our RT-PCR protocol reflects the trends in SARS-CoV-2 variant prevalence over time regardless of sequence mutation. This may aid in the tracking of new variants in the population.

## Introduction

Following the first report of the novel coronavirus, severe acute respiratory syndrome coronavirus 2 (SARS-CoV-2), in 2019 in Wuhan, China (Zhou et al., 2020; Zhu et al., 2020), its rapid spread worldwide has led to the emergence of new lineages and variants, some of which are labeled variants of interest (VOI) or variants of concern (VOC) due to the presence of mutations that alter viral characteristics such as transmissibility and fitness. In Japan, the first report of a patient with coronavirus disease 2019 (COVID-19) was in January 2020 and multiple clusters were identified in the following months. These clusters found in the first months directly descended from the original Wuhan-Hu-1 strain (Sekizuka et al., 2020) and were considered to have originated from the first wave in China, with PANGO lineage B.1.1.162 being prominent in Japan. From March, the number of cases markedly increased worldwide, especially in the US and Europe, and a similar increase in cases in Japan correlated with the outbreak in Europe (Sekizuka et al., 2020). In September 2020, the B.1.1.7 lineage or Alpha variant became prominent in Europe and was later detected in Japan, followed by the detection of the R.1 lineage carrying the E484K mutation (Hirotsu and Omata, 2021; Sekizuka et al., 2021). Recently, the Delta variant, or B.1.617.2, AY.1, AY.2 and AY.3 lineages, has become the most prevalent among positive cases according to the Japanese Ministry of Health, overtaking Alpha and R.1. The Delta variant is known to have increased transmissibility compared with Alpha and is less sensitive to neutralization by antibodies (Campbell et al., 2021; Lucas et al., 2021; Planas et al., 2021), making it a major VOC attenuating the efficacy of currently available vaccines.

This everchanging predominance of variants makes it difficult to screen for SARS-CoV-2 using standard RT-PCR testing because mutations affect primer alignment. We previously verified the use of a set of primers to increase the sensitivity and accuracy of SARS-CoV-2 testing (Tsutae et al., 2020). The JPN-N2 primer set developed by the National Institute for Infectious Diseases, Japan (NIID) (Shirato et al., 2020), and CDC-N1 and CDC-N2 primer sets developed by the US Centers for Disease Control (CDC) (Lu et al., 2020) all detect the nucleocapsid region, but JPN-N2 was reported to have higher sensitivity than CDC-N1 (Tsutae et al., 2020). The fourth primer set included in our screening set, GH-E, detects the envelope region. JPN-N2 is used to surveille positivity in Japan, whereas CDC-N2 is used in the U.S.; however, discrepancy in PCR results has been reported based on which primers are used (Park et al., 2020; Peñarrubia et al., 2020), leading to false-positives or false-negatives (Woloshin et al., 2020). With the continuing increase in the prevalence and emergence of new variants, it is important to assess whether the currently employed testing protocols can accurately detect positive cases. In addition, as the Delta variant is considered to be approximately 60% more infectious than other known variants and has reduced antibody sensitivity (Planas et al., 2021), a testing protocol that reflects the trends in viral load is essential for preventing further infections caused by reduced detection due to mutations affecting RT-PCR.

To date, there have been numerous studies assessing the spread of SARS-CoV-2 and the emergence of new variants internationally, such as in the US and the UK (Bolze et al., 2021; Martin et al., 2021; Volz et al., 2021a, 2021b), but few studies from Japan have examined the recent domestic rapid evolution and variant predominance in detail. We performed next-generation sequencing (NGS) of positive cases of SARS-CoV-2 detected by RT-PCR at the Genesis Institute of Genetic Research, and carried out lineage assessment to detect trends in variant predominance and their correlation with RT-PCR results. A previous study reported a correlation between lower cycle threshold (Ct) values and higher viral load (Corman et al., 2020). In this study, we confirmed a significant correlation between variant prevalence and Ct values, with higher viral loads being observed for Delta cases using all primer sets, and the timing of the increases in cases of different variants in Japan correlated with worldwide trends. This study also demonstrated that our 4-primer set RT-PCR protocol is suitable for the accurate assessment of SARS-CoV-2 positivity regardless of lineage. In addition, we report the first detection of geographically specific AY lineages in Japan.

## Materials and Methods

### Sample collection

Nasopharyngeal swabs and saliva samples for testing were collected at the Genesis Institute of Genetic Research between January and August 2021. Samples were collected from volunteers, testing participants and patients under physician care undergoing clinical testing for SARS-CoV-2 (COVID-19). Samples were anonymized, and laboratory technicians and researchers were blinded to the identity of the patients. Only non-identifying data, such as age, sex and symptoms, were provided. All experimental protocols were approved by the ethics committee of Genesis Healthcare. RNA was extracted using the MGISP-960 system (MGI Tech Co. Ltd.) and RT-PCR was performed using the following primer sets according to the previously reported protocol (Tsutae et al., 2020): JPN-N2, CDC-N1, CDC-N2, GH-E. Primer and probe sequences are listed in Table 1, and binding sites are shown in Fig. 1A. The 730 total samples positive by the JPN-N2 primer set in duplicate or all four primer sets were stored at -80°C until sequencing.

**Table 1.**
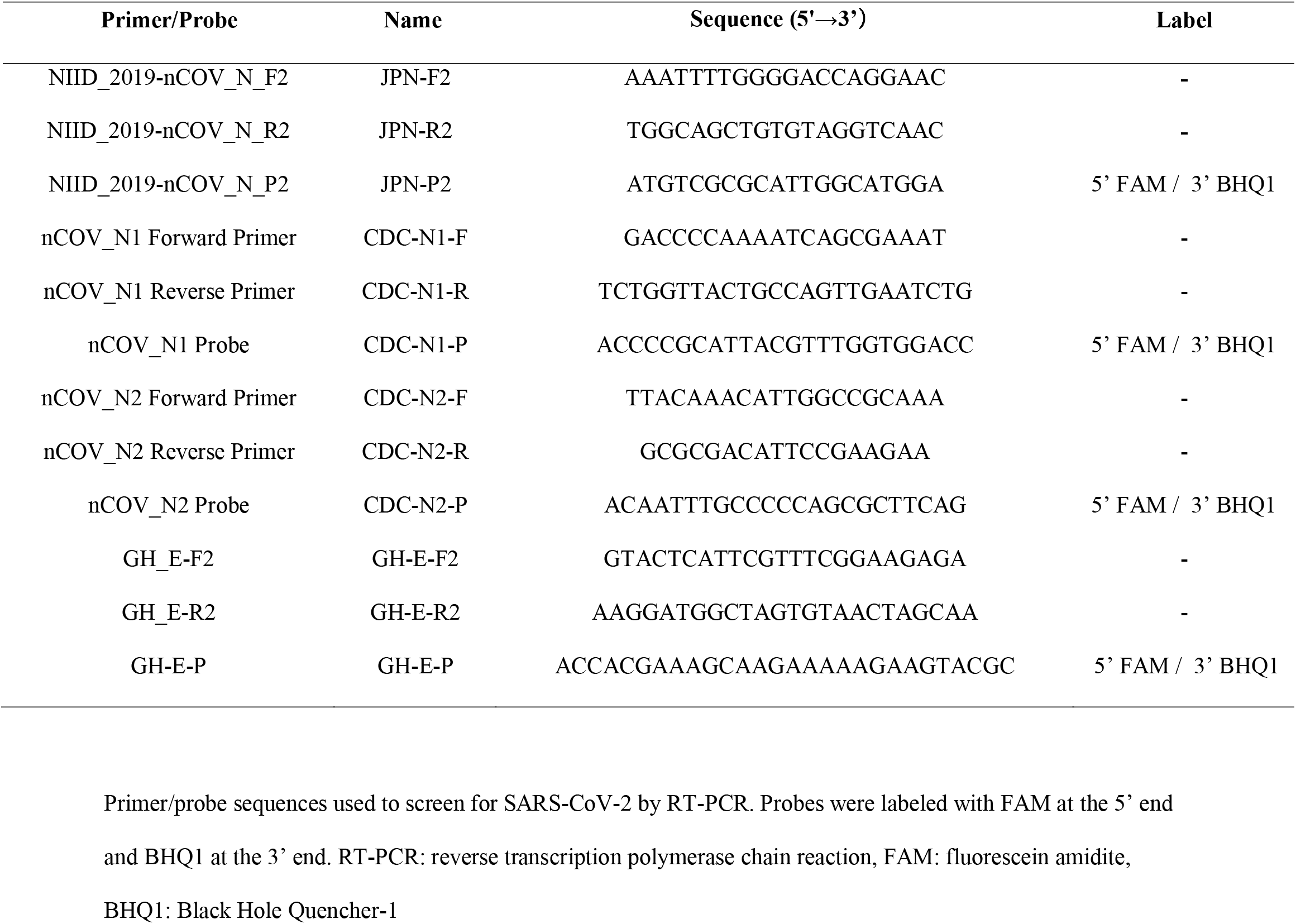
Primer/Probe sets used to detect SARS-CoV-2.

**Figure 1.**
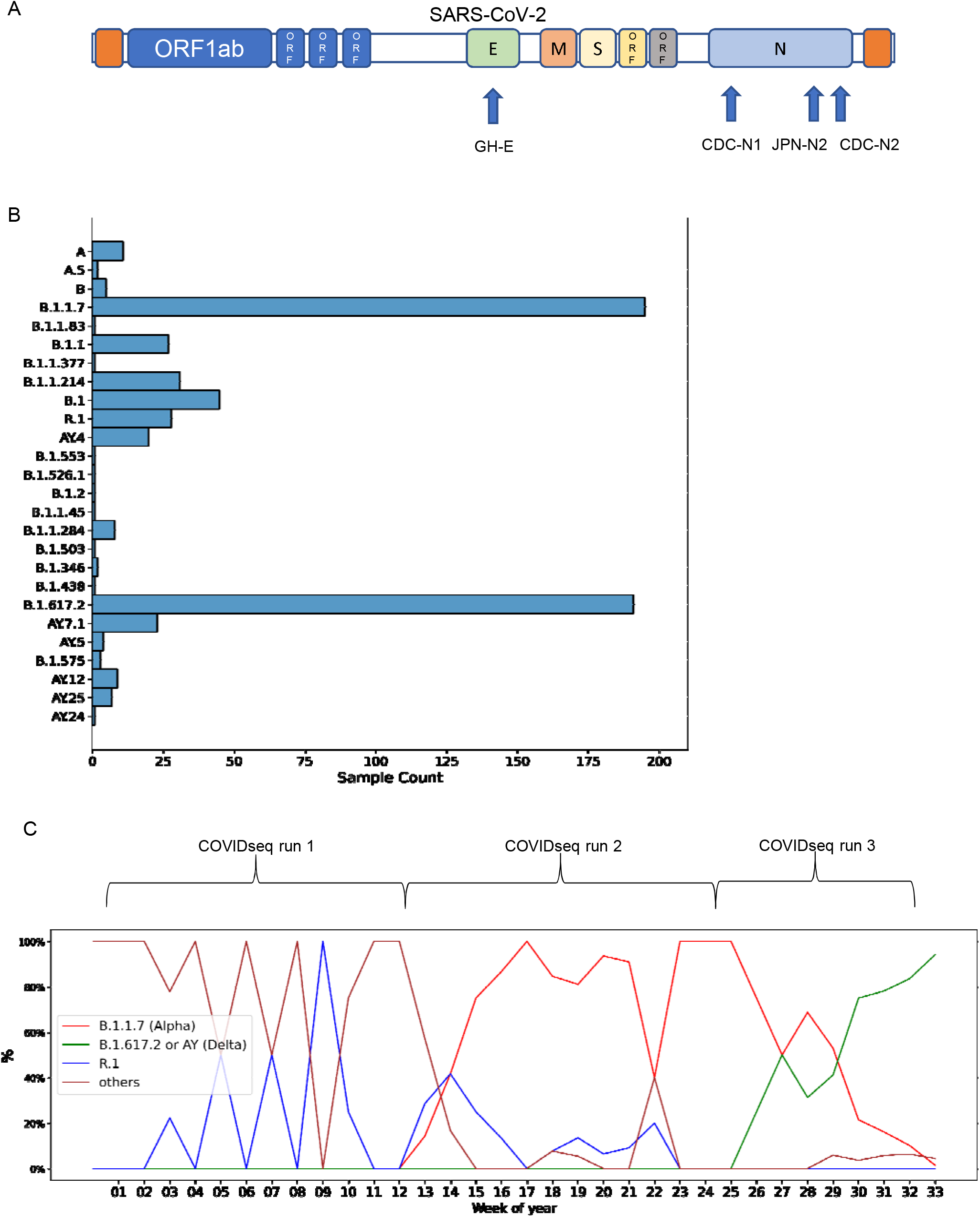
Trends in SARS-CoV-2 lineage prevalence over time. A) Binding sites of each primer set in the SARS-CoV-2 sequence. JPN-N2, CDC-N1 and CDC-N2 sets bind in the nucleocapsid region. The GH-E set binds in the envelope region. B) Detailed lineage breakdown of all collected samples. C) Percentage of detected variants among sequenced samples per week of 2021. Brackets indicate the period during which samples were collected for each COVIDseq run.

### SARS-CoV-2 sequencing (COVIDseq)

SARS-CoV-2 RNA from the 730 collected samples was reverse-transcribed into cDNA, and tagmented and pooled to create libraries following the illumina COVIDSeq Test Reference Guide. Samples were sequenced using the illumina NovaSeq 6000 system SP flow cell, comprising the NovaSeq 6000 SP reagent kit v.1.5 and NovaSeq Xp 2-lane kit v.1.5, with 100 cycles for the first run, 200 cycles for the second run and 200 cycles for the third run. The resultant sequence data in the SampleSheet.csv file were corrected strictly following the SampleSheet COVIDSeq protocol (‘illumina-adapter-sequences-1000000002694-15.pdf’) and uploaded onto the illumina DRAGEN Bio-IT Platform. The illumina DRAGEN COVIDSeq Test (RUO) App was used for consensus sequence calling and a consensus genome in FASTA format was generated for samples with a result of “SARS-CoV-2 Detected” (for details, see ‘dragen-covidseq-test-ruo-v130-app-guide-1000000129548-01.pdf’). These FASTA files were next input into the illumina DRAGEN COVID Lineage App. The NextClade tool (Nextstrain, Nextclade: https://clades.nextstrain.org/) was used for clade assignment and phylogenetic analyses, and Pangolin (https://cov-lineages.org/; corresponding GitHub repository: https://github.com/cov-lineages) was used for lineage calling (For more information, see https://www.illumina.com/products/by-type/informatics-products/basespace-sequence-hub/apps/dragen-covid-lineage.html). Sequence quality control (QC) parameters included missing data, mixed sites, private mutations and mutation clusters. Only samples that passed QC for lineage calling and clade assessment were used in subsequent analyses.

### Statistical analysis

Ct values for each primer set among variants were compared using the Kruskal-Wallis H test (one-way non-parametric ANOVA) followed by post hoc Conover’s test. Values are presented as the mean ± standard error. Differences between the expected and observed frequencies of symptoms among the Alpha, Delta and ‘others’ variants were assessed using the chi-squared test. A p-value <0.05 was considered significant.

## Results

### Trends in SARS-CoV-2 infections during 2021 in Japan based on NGS

From January 2021, the R.1 lineage was detected in up to approximately 38% of positive cases in Japan after its first report in November 2020 (Hirotsu and Omata, 2021). As R.1 was first detected in the U.S. a month earlier, this change in R.1 prevalence in Japan and other countries was similar (Hirotsu and Omata, 2021). In addition, an increase in B.1.1.7 or Alpha cases was reported in March 2021 during the fourth wave and B.1.617 or Delta was first detected in April according to the Japanese Ministry of Health. As we perform routine commercial testing of SARS-CoV-2, we hypothesized that our positive cases reflect the prevalent variants during a given period. We therefore performed NGS of positive samples received for SARS-CoV-2 screening at the Genesis Institute of Genetic Research to identify the viral lineages causing infection in the Japanese public in 2021. Three separate COVIDseq runs were conducted using samples that were positive for SARS-CoV-2 by all 4 primer sets shown in Fig. 1A collected from January to April in the 1^st^ run, April to June in the 2^nd^, and July to August in the 3^rd^. In total, 730 samples were sequenced. After QC assessment for lineage calling and clade assignment, 620 samples were used for analysis. Of the 620 samples that passed QC, 190 were tested only using the JPN-N2 primer set in duplicate and 430 samples were tested using the four-primer set and were positive with all four sets. Based on phylogenetic clade assignment of known VOC and VOI, 195 samples were determined to be Alpha, 28 were R.1, 255 were Delta and 142 were other lineages (Fig. 1B, Supp. Figs. 1, 2, 3). Alpha accounted for 47% of cases collected between January and June, and 10% of cases during this period were R.1 (Fig. 1C). However, in the 3^rd^ run using samples from July to August, only 17% of cases were Alpha and no cases of R.1 were detected, suggesting that these lineages were overtaken, most likely by Delta. Delta was first detected in our samples in the end of June and accounted for 78% of cases in the 3^rd^ run, demonstrating a rapid increase in prevalence (Fig. 1C). Delta lineages were separated into B.1.617.2 and AY.4-AY.25, with the majority being the former (Fig. 1C). This suggested that the different variants displaced each other within a short period of around two months according to each wave of infection.

### SARS-CoV-2 VOC are associated with a higher frequency of symptoms

It is well known that viral mutations can cause more severe cases of COVID-19 (Hahn et al., 2020; Nagy et al., 2021; Voss et al., 2021). We thus analyzed the reported symptoms and compared their frequencies between each VOC and the “other” lineages. At the time of sampling, the presence of the following symptoms was asked: fever, tiredness, chills, headache, muscle pain, dyspnea, cough, sore throat, runny nose, abnormal olfaction/taste, diarrhea, nausea, and rash. The most commonly reported symptom in R.1 and ‘others’ cases was cough in approximately 20% and 15%, respectively, whereas that in Alpha and Delta cases was fever in approximately 24% and 34%, respectively (Fig. 2). Consistent with the reports of more severe cases with VOC infection, Alpha and Delta cases had a significantly higher rate of fever and cough than ‘others’ cases (p <0.05). Furthermore, headache and tiredness were significantly more frequently reported in Alpha and Delta cases (p <0.05), suggesting an increase in severity. Of note, approximately 15% of ‘others’ cases reported abnormal olfaction/taste, which is often one of the first symptoms noted in mild COVID-19 (Spinato et al., 2020). However, it was reported slightly less in Delta and Alpha cases (Fig. 2). Muscle pain and dyspnea were also more common in Delta and Alpha cases, although not significantly.

**Figure 2.**
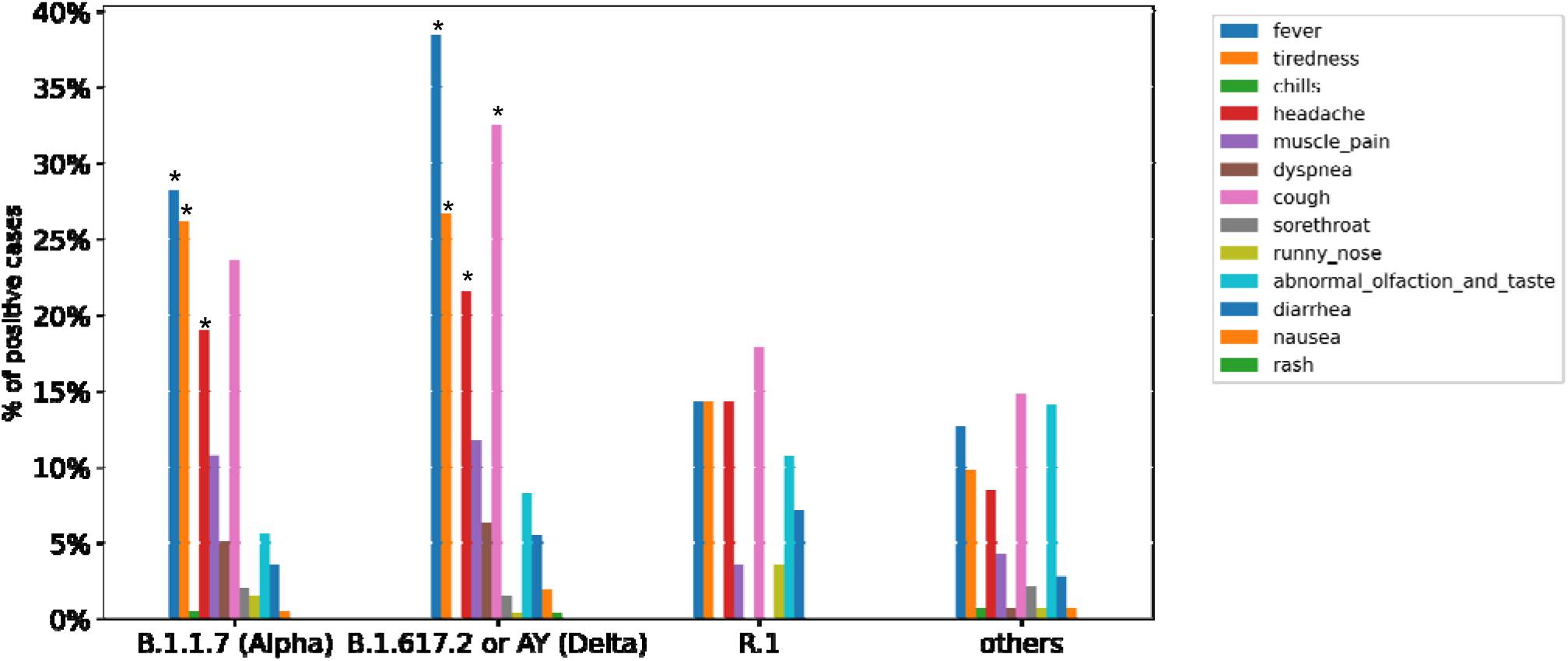
Reported symptoms associated with each variant. Frequencies of reported symptoms in cases of each variant. Differences were assessed by the chi-squared test. * indicates significantly more often compared with the ‘others’ group.

We next examined the relationship between Ct values and the number of reported symptoms at the time of sampling, but no significant trend was observed in our data (Supp. Fig. 4).

Based on these results, VOC may cause higher rates of commonly observed symptoms that are not specific to SARS-CoV-2 infection.

### Increased viral load is associated with the increased prevalence of Delta cases

The rapid spread of VOC is considered to be due to an increased viral load via faster replication rates (Campbell et al., 2021; Li et al., 2021; Volz et al., 2021a). Alpha was reported to have higher transmissibility than non-VOC and Delta was reported to have even higher transmissibility due to an increased viral load (Li et al., 2021; Wintersdorff et al., 2021). Consistent with this, we noted a significant difference in Ct values with all RT-PCR primer sets among variants. With the JPN-N2 primer set, the mean Ct value for ‘others” lineages was 30.1±0.3, that for R.1 was 27.5±1.1, that for Alpha was 25.9±0.3 and that for Delta was 24.4±0.3 (Fig. 3A). With the CDC-N1 primer set, the mean Ct values were 29.6±0.5, 27.2±1.1, 25.4±0.4 and 23.7±0.4, respectively (Fig. 3B). With the CDC-N2 primer set, the mean Ct values were 30.1±0.5, 27.1±1.1, 25.8±0.4 and 23.9±0.4, respectively (Fig. 3C). Similarly, with the GH-E primer set, the mean Ct values were 30.9±0.5, 26.8±1.2, 26.9±0.4 and 25.3±0.4, respectively (Fig. 3D). Each variant had significantly lower Ct values than the ‘others’, with Delta having the lowest Ct values among all variants, consistent with its higher viral load and infectivity. As it was previously reported that Ct values >34 are the limit for viral isolation from samples (La Scola et al., 2020), our samples were from contagious individuals.

**Figure 3.**
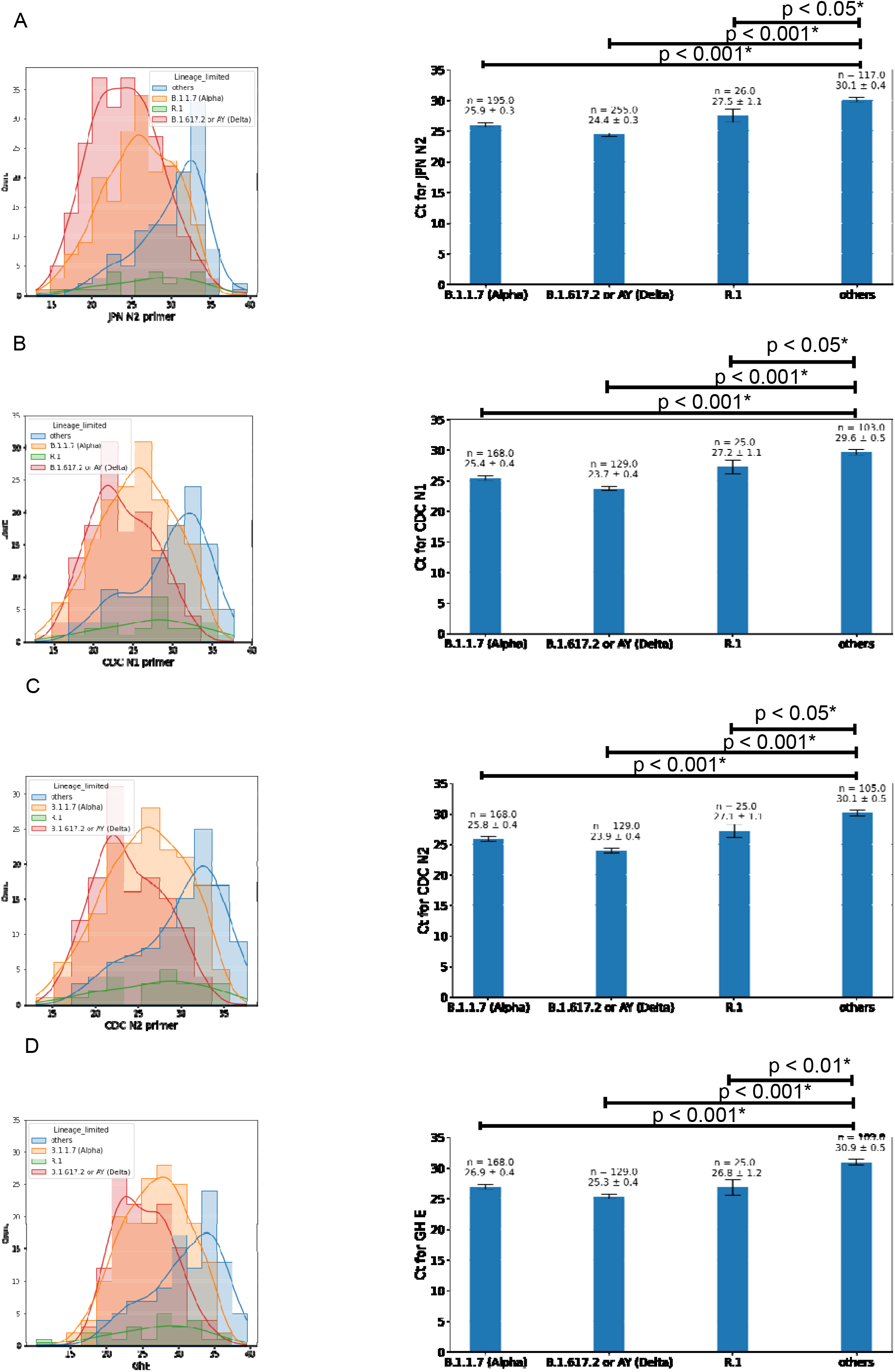
Differences in Ct values among variants by RT-PCR primer set. A) Ct values with the JPN-N2 primer set for each variant. B) The mean ± standard error Ct value for each lineage with the JPN-N2 primer set. C) Ct values with the CDC-N1 primer set for each variant. D) The mean ± standard error Ct value for each lineage with the CDC-N1 primer set. E) Ct values with the CDC-N2 primer set for each variant. F) The mean ± standard error Ct value for each lineage with the CDC-N2 primer set. G) Ct values with the GH-E primer set for each variant. H) The mean ± standard error Ct value for each lineage with the GH-E primer set. Ct values were compared among variants using the Kruskal-Wallis H test (one-way non-parametric ANOVA) followed by post hoc Conover’s test. P-values with an * indicate significantly different from the ‘others’ group.

This decrease in Ct correlated with the trends in viral prevalence over time, demonstrating a decline with each successive wave of infection spurred by the introduction of each VOC. As shown in Fig. 4A, the mean Ct using our 4-primer set screening method ranged between approximately 28 and 35 from January to the end of February, during which ‘others’ and R.1 lineages comprised the majority of cases (Figs. 1C, 4A). Coinciding with the spread of Alpha in the fourth wave, the mean Ct markedly decreased to around 24-28 during March and April, indicating its higher infectivity. From the last week of May to June, there was a temporary drop in Alpha cases, and the mean Ct increased back to around 30 accordingly. Then, from the first week of June until August, the mean Ct values further dropped to around 24-26 in parallel with the introduction and spread of Delta (Figs. 1C, 4A).

**Figure 4.**
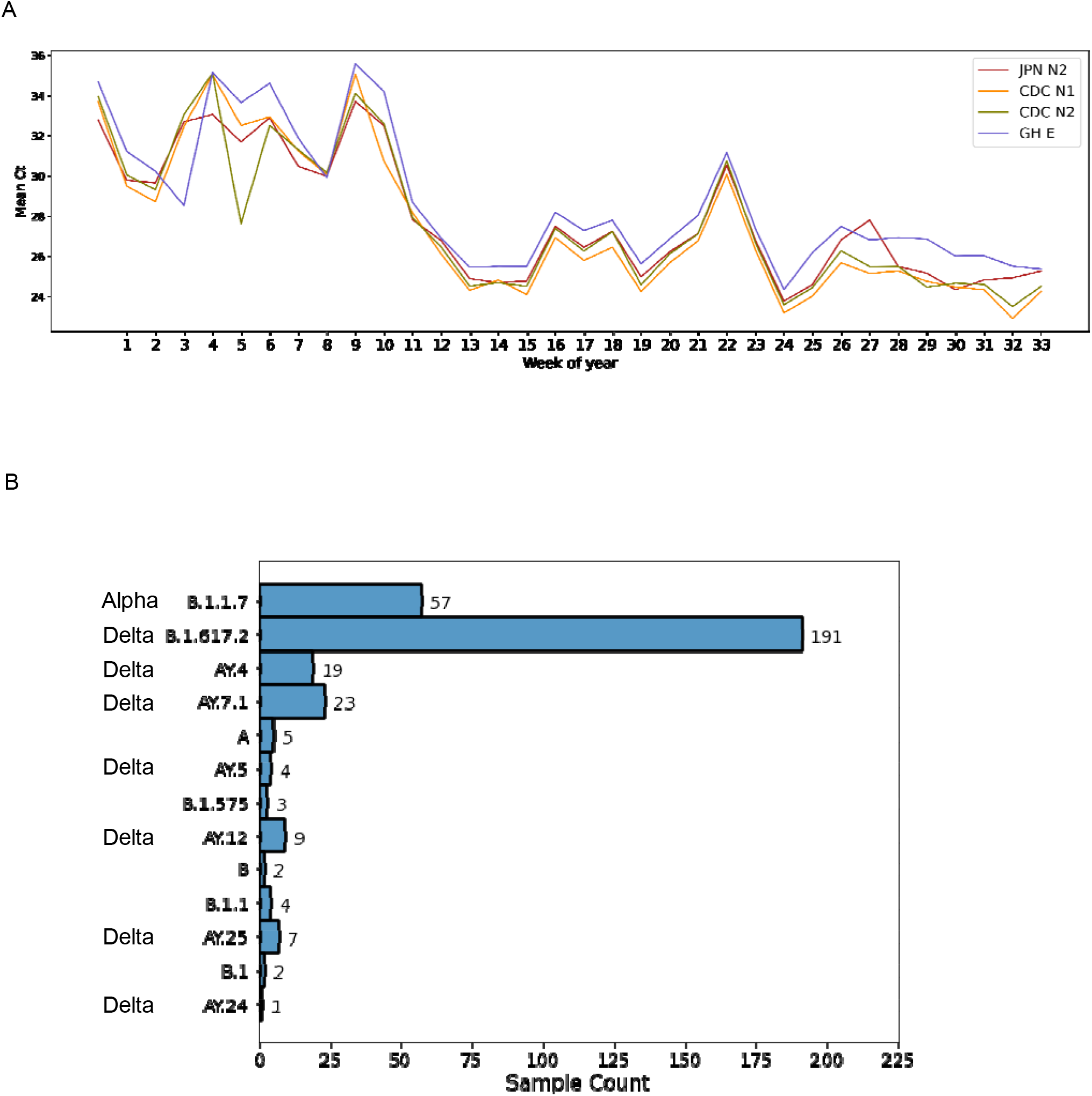
Correlation between SARS-CoV-2 variants and RT-PCR Ct values. A) Graph of the mean Ct value with each primer set for each variant over time. B) Detailed breakdown of lineages detected during COVIDseq run 3.

To confirm that the changes in Ct values were due to increased viral load rather than mutations affecting primer binding, we examined COVIDseq data for any mutations in the primer binding sites. Among the 4 primer sets, only 3 of the 620 sequenced samples had a mutation at primer binding sites (Table 2). Two of these samples were Delta and one was Alpha, but only one of these mutations, specifically G29179T in the CDC-N2 forward primer binding site, was considered to reduce binding. This confirmed that the lower Ct values in VOC cases were due to increased viral load. As viral loads in Delta cases were reported to be 1000-times higher than those in non-VOC cases (Li et al., 2021), the Ct values observed using our RT-PCR method reflected the real-time trends in the prevalence of VOC.

**Table 2.** Mutations in primer binding sites.

| Sample | Clade | Changes in primer site | JPN-N2 | CDC-N1 | CDC-N2 | GH-E |
| --- | --- | --- | --- | --- | --- | --- |
| 20210827_A113 | 21A<br>(Delta) | USCDC_N1_F:C28298A | 29.49 | 28.61 | 28.84 | 29.58 |
| 20210827_A222 | 21A<br>(Delta) | GHE_F:G26272T | 20.51 |  |  |  |
| 20210712_A135 | 20I<br>(Alpha,<br>V1) | USCDC_N2_F:G29179T | 27.63 | 26.76 | 31.39 | 22.65 |

Mutations identified in primer binding sites by COVIDseq. Listed Ct values are those from the initial SARS-CoV-2 RT-PCR screening.

### Delta displaced Alpha as the major variant in Japan within 1 month

Although travel restrictions, quarantine and vaccination measures were implemented worldwide, Delta rapidly established itself as the dominant variant in countries like India, the UK, the Netherlands and Australia, (Campbell et al., 2021; Wintersdorff et al., 2021) and displaced Alpha in the US by July 2021 (Bolze et al., 2021). Alpha was the dominant VOC detected in our samples from March to June (Figs. 1C, 4A). The first reported case of Delta in Japan was in April 2021, whereas we first detected the Delta variant, specifically B.1.617.2, in mid-June in a sample from Chiba prefecture (Figs. 1C, 4A). By mid-July, Delta accounted for 50% of all positive samples sequenced and its prevalence continued to increase to nearly 90% into August, overtaking Alpha (Figs. 1C, 4A). Therefore, although expected based on previous worldwide surveillance and its increased transmission rate, our sequencing results confirmed that the Delta variant rapidly dominated Japan.

### Detection of country-specific Delta lineages in Japan

In addition to B.1.617.2, we detected Delta lineages AY.4-AY.25 (Fig. 4B), which are subclades within B.1.617.2 that exhibit significant geographic clustering, although there is no functional biological difference. As of August 21, 2021, AY.1-AY.25 lineages are included in the pangoLEARN model, with lineages AY.4-AY.11 being listed as predominant in the UK. During our third sequencing run using samples collected in July and August, we detected 23 cases of the Delta lineage AY.7.1, which is specific to Denmark. There were also 19 cases of AY.4, 4 cases of AY.5, 9 cases of AY.12, 7 cases of AY.25 and 1 case of AY.24 (Fig. 4B). These lineages are most prominent in the UK, Israel, the US and Indonesia, respectively (O’Toole et al., 2021). Considering their geographical diversity, these lineages may have been introduced into Japan through international travel from July 2021, but this point requires further epidemiological study.

## Discussion

During 2020, the first (March to April 2020), second (July to September 2020), and third (October 2020 to January 2021) waves of COVID-19 in Japan were characterized by B.1.1.162, B.1.1.284, and B.1.1.214 lineages, respectively (Sekizuka et al., 2020). According to a previous study, variants of SARS-CoV-2 evolve through natural selection and dominate the population (Hou et al., 2020). NGS analysis has made it possible to trace the spread of SARS-CoV-2 and identify specific lineages infecting a population. This is particularly important as the emergence of mutations has led to the rapid spread of VOC and VOI that may be less sensitive to available vaccines. We used Ct values from RT-PCR and NGS data to investigate the trends in variant prevalence in Japan based on samples collected from January to August 2021. The most prevalent strain changed within an approximately two-month period between sequencing runs. From the period of January to March, ‘others’ and R.1 lineages were most prevalent among our samples. R.1 was first reported in the US in October 2020 and in Japan in November, but the reported isolates where phylogenetically similar; therefore, it is possible that the Japanese isolate originated from the US isolate (Sekizuka et al., 2020). By mid-April 2021, nearly 40% of the R.1 lineage entries in the Global Initiative on Sharing All Influenza Data (GISAID) database were from Japan (Hirotsu and Omata, 2021). We observed the highest number of cases at the end of March and into April, which was consistent with the peak incidence in Japan and the US according to international lineage tracking at cov-lineages.org (O’Toole et al., 2021).

However, despite the rapid spread of R.1, it was quickly dominated by the more transmissive Alpha in Japan and overseas (Washington et al., 2021). We first noted a significant drop in Ct values mid-March, which coincided not only with an increase in Alpha cases detected by our sequencing, but also with the worldwide increase based on lineage tracking. The use of Ct values to assess mortality, prognosis, disease severity and infectivity has been reviewed in many previous studies (Rabaan et al., 2021; Rao et al., 2020), but it is well known that they are affected by many factors, including primer design. Our RT-PCR assay uses 4 primer sets, 3 of which detect the nucleocapsid region, in order to increase the detection sensitivity and specificity while using the same PCR program. We previously reported that the CDC-N1 primer set is prone to false-positives, and should not be used alone without CDC-N2 or JPN-N2, which have a higher accuracy in the Japanese population (Tsutae et al., 2020). In addition, among all 620 sequenced samples, only one Alpha case carried a mutation that may have reduced CDC-N2 primer binding, leading to a single higher Ct value (Table 2). The overall observed changes in Ct values with all primer sets were congruent with changes in the lineages isolated over time, with lower Ct values correlating with the increase in cases of more infective Alpha and Delta variants, indicating that the varying mutations in these lineages did not significantly affect our RT-PCR. Of note, we only observed a significant correlation between the number of symptoms and presence of VOC, but not between Ct values and symptoms. This is consistent with a study that reported similar Ct values between asymptomatic and preymptomatic cases (Singanayagam et al., 2020), as well as a study in the UK stating that Ct values ultimately depend on the virus strain in the population (Walker et al., 2021). Therefore, our study revealed that Ct values of multiple primer sets can be a good reflection of the transmissibility of the most prominent SARS-CoV-2 lineage in a population at a given time. Although our study has the limitations of symptom onset data being unavailable and the timing of sample collection varying throughout the course of infection, it suggested that a rapid decrease in Ct values in routine testing indicates the introduction of a new, more infective variant, whereas an increase may indicate its fading out from a population.

The R.1 variant, which was reported to have potential escape mutations in the N-terminal domain (W152L) and spike protein (E484K) (Hirotsu and Omata, 2021), was recently re-detected in the US amidst the rampant spread of Delta (Cavanaugh et al., 2021). This variant was not detected in our samples after June, suggesting that variants can reemerge in different populations worldwide. Variants harboring E484K, L452R, or N501Y are of particular concern because they are related to escape from antibody neutralization (Cherian et al., 2021; Lucas et al., 2021). As such, even after vaccination or previous infection, if these variants are reintroduced to a population, they may become dominant again. As nearly 80% of the Japanese population has received two doses of the SARS-CoV-2 vaccine as of February 2022, it will be of interest to investigate how the presence of antibodies affects the average Ct values and transmissibility of known and new variants such as Omicron. Furthermore, as the third vaccine is just now being administered in Japan at present, it is important to identify mutations in highly transmissible strains in order to prevent the spread of even more infectious variants that escape antibody neutralization. Our NGS data will be highly useful for investigating which mutations were conserved in the SARS-CoV-2 sequence throughout the rapid fluctuation of lineages present in Japan, aiding in the estimation of future VOC through machine learning.

## Data Availability

NGS data from the first and second COVIDseq runs were uploaded to GISAID.

## Ethics Statement

Samples were collected from volunteers, testing participants and patients under physician care undergoing clinical testing for SARS-CoV-2 (COVID-19). Samples were anonymized, and laboratory technicians and researchers were blinded to the identity of the patients. Only non-identifying data, such as age, sex and symptoms, were provided. All experimental protocols were approved by the ethics committee of Genesis Healthcare.

## Data availability

NGS data from the first and second COVIDseq runs were uploaded to GISAID.

## Acknowledgements

None.

## Author Contributions

I.S.B. conceived the study. D.W., J.T., and H.A. processed samples and performed RT-PCR. D.W, M.O., S.N., E.S., and J.T. performed next-generation sequencing. C.C. and W.O. carried out data analysis and lineage assignment. D.W. wrote the manuscript. All authors reviewed the data and approved the manuscript.

---

**Supplementary Figure 1. Phylogenetic tree for COVIDseq run 1**

Phylogenetic tree for samples collected between January and April 2021 generated by https://clades.nextstrain.org/tree.

**Supplementary Figure 2. Phylogenetic tree for COVIDseq run 2**

Phylogenetic tree for samples collected between April and June 2021 generated by https://clades.nextstrain.org/tree.

**Supplementary Figure 3. Phylogenetic tree for COVIDseq run 3**

Phylogenetic tree for samples collected between June and August 2021 generated by https://clades.nextstrain.org/tree.

**Supplementary Figure 4. Plot of the number of symptoms at sampling versus Ct values**

Graph of the average number of symptoms reported at the time of sampling versus rounded Ct value. No significant trend was observed.

