## Supplementary figures and images for "Rapid displacement of SARS-CoV-2 variants within Japan correlates with cycle threshold values on routine RT-PCR testing"

### Supplemental Figures

#### Slide 1
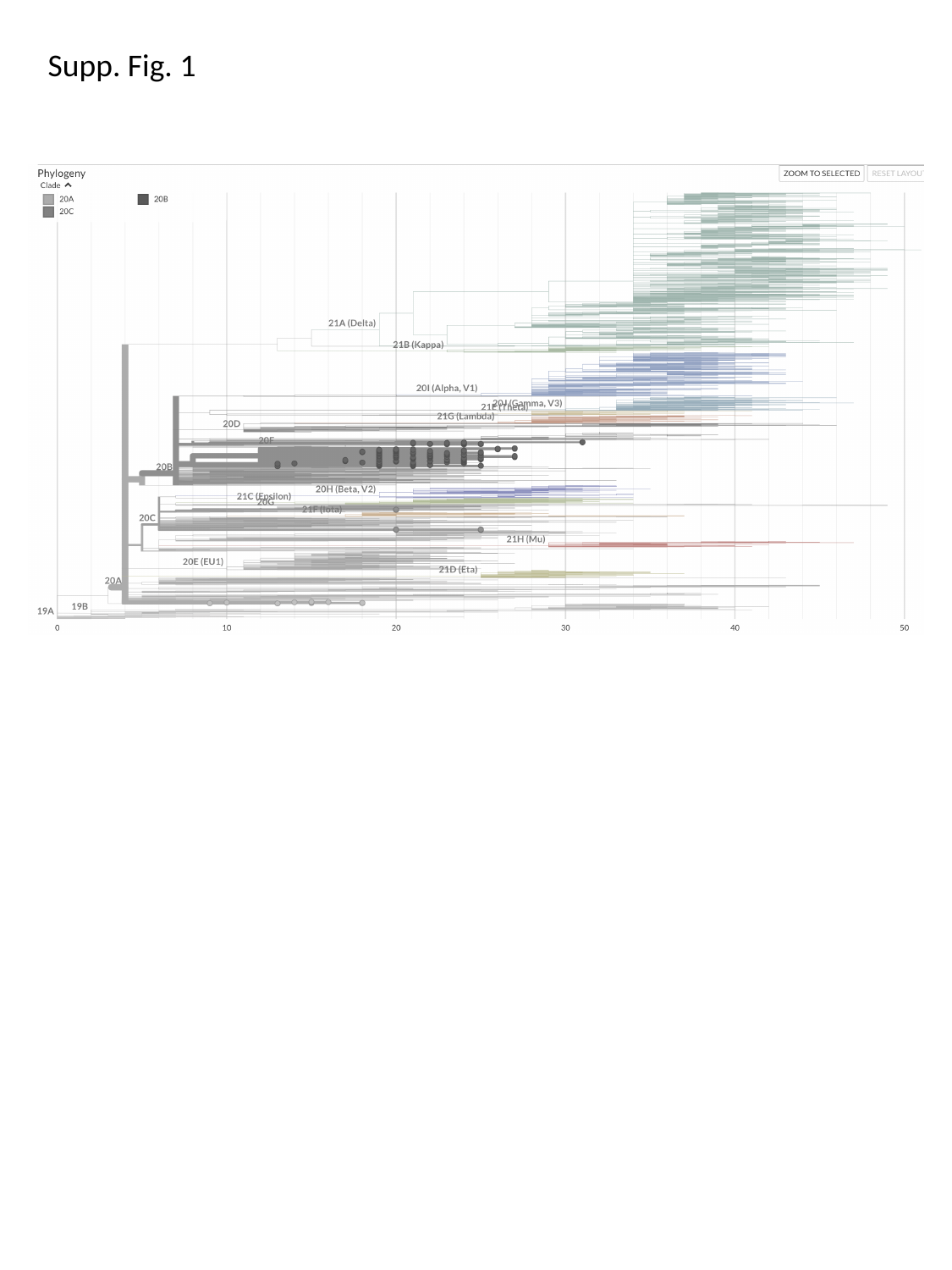

Supp. Fig. 1

#### Slide 2
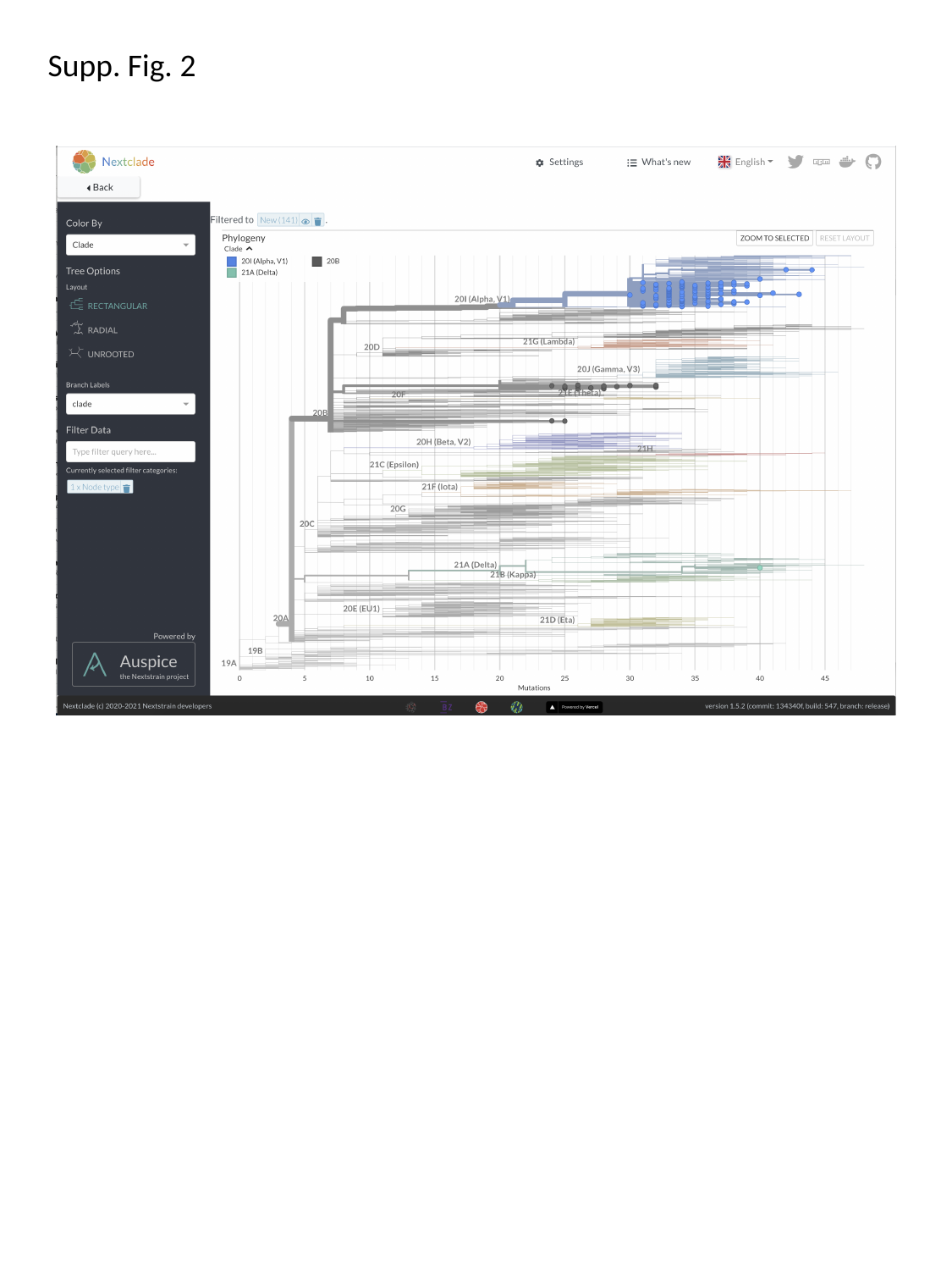

Supp. Fig. 2

#### Slide 3
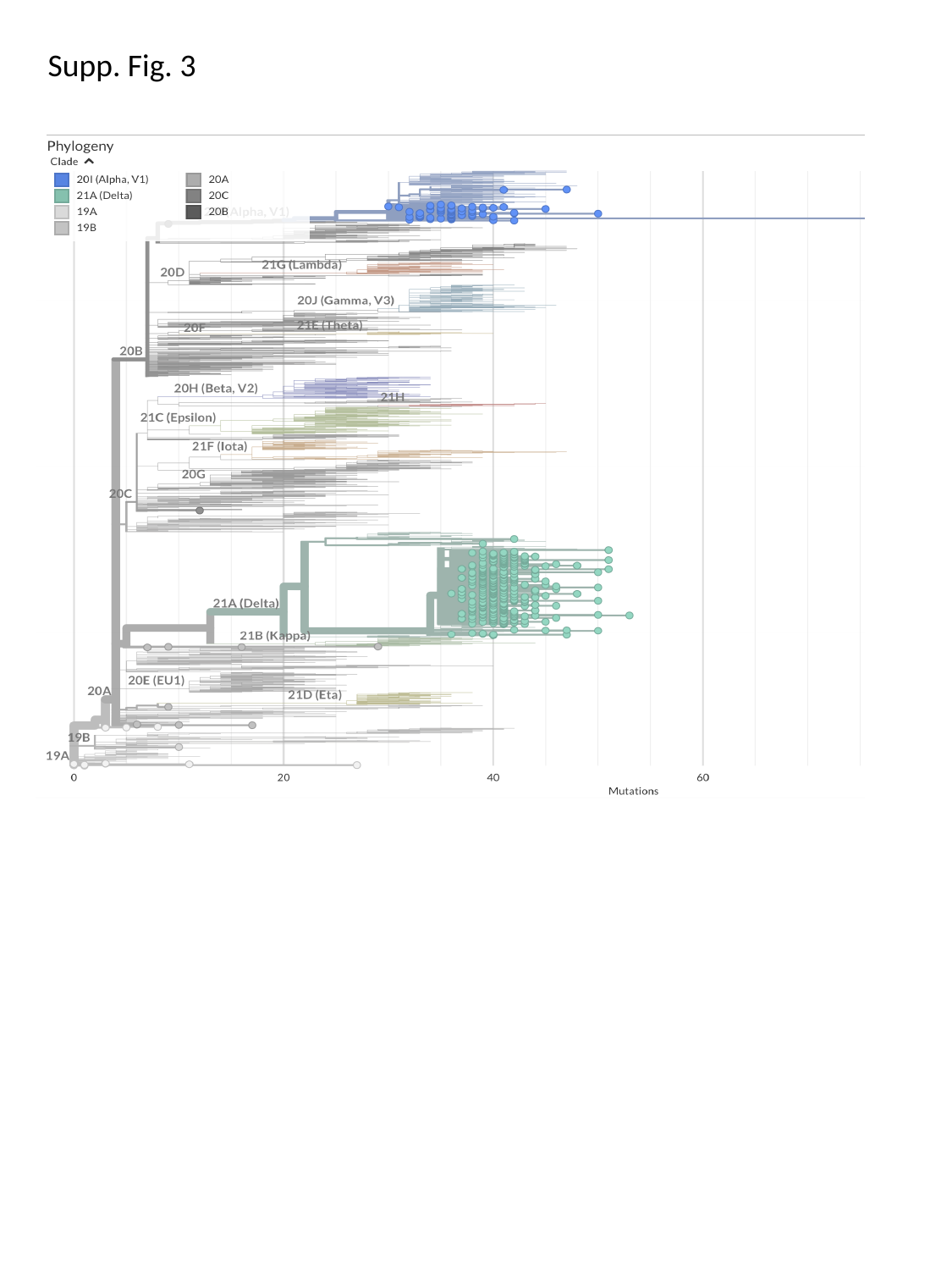

Supp. Fig. 3

#### Slide 4
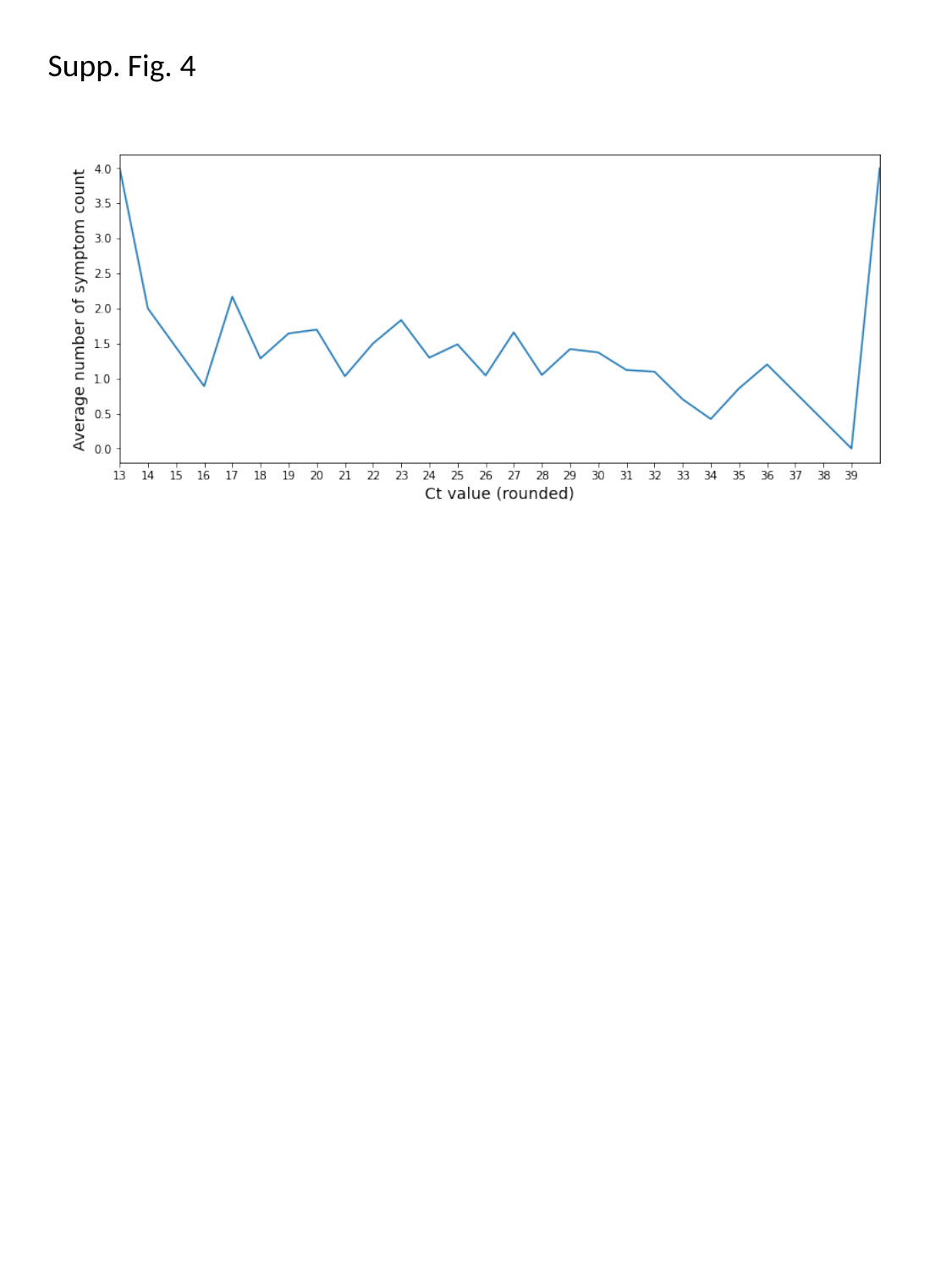

Supp. Fig. 4
